# Industry differences in managers’ experiences of work capacity in employees with common mental disorders: a cross-sectional study on the Swedish labour market

**DOI:** 10.1101/2024.09.06.24313226

**Authors:** Lisa Björk, Jenny Hultquist, Gunnel Hensing, Monica Bertilsson

## Abstract

The aim of this study was to assess industry differences in managers’ experiences and ratings of work capacity in employees with common mental disorders (CMDs). Swedish managers (N=1819) were grouped into three industry classifications. Differences between industries in managers’ experiences were investigated by chi-squared tests. The managers’ ratings of how work capacity was affected by CMDs were analysed using MANCOVA, adjusted for organizational size and managers’ span of control. The proportion of managers who had experienced several employees with CMDs was higher in municipalities and counties, and in pink-collar work, education, health and social care settings compared with other industries. There was no significant effect of industry on managers’ ratings of how work capacity may be affected by CMDs. Contrary to assumptions, there were no differences in how managers perceived capacity to work between industries. Therefore, it is more urgent that managers receive the support they need to handle employees with reduced work capacity due to CMDs rather than to tailor such support to different industries.

## Introduction

Employers are responsible for workers’ health and safety. This responsibility is executed by managers through universal preventive efforts regarding the work environment and through individual support to workers in vocational rehabilitation [1–5]. In particular, managers highlight vocational support as challenging in the case of CMDs [6].

The World Health Organization has estimated the prevalence of mental disorders to be approximately 15% among working adults [7, 8]; depression and anxiety are the most widespread disorders. Most people with these disorders remain at work, but others experience decreased work capacity and become sick-listed. In Sweden, sickness absence (SA) with psychiatric disorders tends to be longer than in other diagnostic groups, and recurrence of sickness absence is higher [9–11]. Length of SA and recurrence is related to the severity of the disorder but also to the adaptability of the work situation. Thus, work accommodation is important to prevent SA but also for vocational rehabilitation [5]. However, qualitative studies have shown that managers find it difficult to accommodate work for employees with CMDs [6, 12]. This is not surprising because work capacity in relation to CMDs is a complex phenomenon [13]. In their review, Lederer et al. [14] concluded that work capacity is determined by individual characteristics as well as by dimensions in the environment. Work capacity has been described as the result of a dynamic interaction between the individual, the work environment and the work tasks [15]. In earlier qualitative studies of people with lived experiences, work capacity is described as multifaceted and not limited to paid work only [16, 17]. To reach the optimal level of an individual’s work capacity, work adjustments are often necessary in terms of changes in work tasks and the work environment, or both.

In the present study, we investigate whether managers in different industrial sectors perceive work capacity in employees with CMDs differently. This approach was based on the uneven distribution of SA across the labour market. Most prominent are the differences between female- and male-dominated sectors, with higher CMD-induced SA levels in both women and men in the female-dominated sectors. We assumed that the varying characteristics of different industries may affect work capacity of employees with CMD differently. If so, managers in different industries might encounter dissimilar aspects of a reduced work capacity among their employees. As an example, managers in the health care sector might notice reductions in social capacity, whereas managers in the IT sector might encounter reductions in cognitive capacities.

Given the uneven distribution of SA across industries, the key role of managers in vocational rehabilitation and the lack of studies in this field, the aim of this study is to assess industry differences in managers’ experiences and ratings of work capacity in employees with CMDs. More specifically, we analyse two research questions:

Q1 What proportion of managers had experienced one or more employees with a CMD during the last 2 years in different industrial sectors categorized by (i) ownership, (ii) work object and (iii) work object + gender composition?
Q2 Are there any differences in managers’ ratings of how work capacity is affected in employees with CMDs in different industrial sectors categorized by (i) ownership, (ii) work object and (iii) work object + gender composition?

## Materials and methods

This explorative, cross-sectional study is a part of the New Ways - Mental Health at Work research programme and the specific project “Managers’ Perspective - A Missing Piece”. The project focuses on Swedish managers’ attitudes towards knowledge and experiences of employees with CMDs. This study was designed as a cross-sectional survey study, with data collected between June 14 and August 8 in 2017.

### Sample

Participants were recruited from the Swedish Citizen Panel at the SOM Institute, University of Gothenburg, and through the HELIX Competence Centre at Linköping University, Sweden. The Citizen Panel consists of self-recruited participants and the HELIX Centre is a collaboration between 22 private and public organizations. Identification of managers in the Citizen Panel was done using two questions on managerial position included in the 26th panel survey in 2017 [18]. Five thousand managers were invited to participate in the study (Fig 1). The HELIX Competence Centre provided additional email addresses for managers (n=556) from eight of the collaborating organizations. The response rate was 71%. The final sample in this study consisted of 1819 managers who had experienced at least one employee with a CMD during the last 2 years. The questionnaire was pilot-tested on ten managers before distribution.

**Fig 1.**
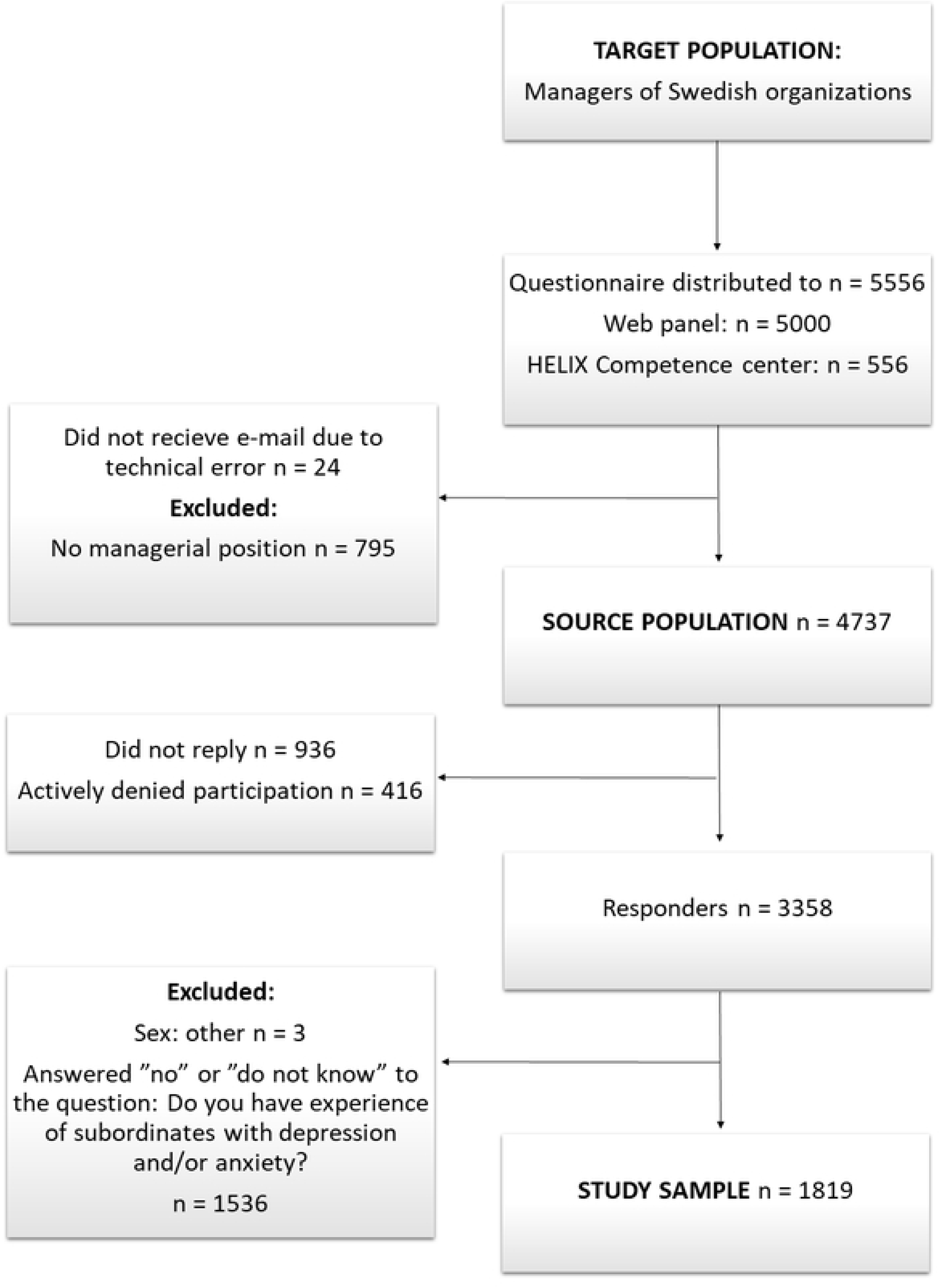
Flowchart of the sampling process.

### Measures

#### Managers’ experience of employees with CMDs

To measure managers’ experience of employees with CMDs, the participants were asked: “In the past two years, have you had employees at your current workplace who have had depression and/or anxiety disorder?”. Of the 1819 managers included in this study, 927 (51%) answered “Yes, one staff member” and 892 (49%) “Yes, several staff members”.

#### Managers’ experience of work capacity in employees with CMDs

The managers were also asked: “Think back to those employees who have had depression and/or anxiety disorders in the past two years at your current workplace. Based on your opinion, how was the work capacity of these employees affected?”. This question was followed by nine items on how work capacity can be affected (Table 1). The nine items were derived from an instrument measuring work capacity in workers with CMD [19]. The nine items constitute two indices: task-oriented work capacity (Cronbach’s alpha, 0.81) and relational work capacity (Cronbach’s alpha, 0.70). Construction of the indices was preceded by a principal component factor analysis (see S1 Table).

**Table 1.** Swedish managers’ ratings of how they thought CMDs affected work capacity in their employees: frequencies and proportions from a cross-sectional web survey, 2017.

| How was the employee’s work capacity affected regarding: | Answer options, n (%) |  |  |  |  | Total, n (%) |
| --- | --- | --- | --- | --- | --- | --- |
|  | Not affected at all | Became somewhat more difficult | Became more difficult | Became much more difficult | Do not know |  |
| Task-oriented work capacity |  |  |  |  |  |  |
| Getting started on work assignments | 59 (3.3) | 436 (24.4) | 620 (34.7) | 591 (33.1) | 81 (4.6) | 1787 (100) |
| Being able to prioritize among work tasks | 41 (2.3) | 262 (14.7) | 495 (27.7) | 909 (50.9) | 80 (4.5) | 1787 (100) |
| Being able to concentrate | 26 (1.5) | 276 (15.4) | 583 (32.6) | 799 (44.7) | 103 (5.8) | 1787 (100) |
| Remembering | 79 (4.4) | 395 (22.1) | 561 (31.4) | 556 (31.1) | 195 (10.9) | 1786 (100) |
| Maintaining the work pace | 54 (3.0) | 376 (21.0) | 576 (32.2) | 724 (40.5) | 57 (3.2) | 1787 (100) |
| Relational work capacity |  |  |  |  |  |  |
| Interacting with other people | 79 (4.4) | 553 (31.0) | 576 (32.3) | 541 (30.3) | 37 (2.1) | 1786 (100) |
| Participating in social work environments (such as coffee breaks) | 187 (10.5) | 551 (30.8) | 475 (26.6) | 461 (25.8) | 113 (6.3) | 1787 (100) |
| Keeping calm and not getting upset | 130 (7.3) | 415 (23.2) | 479 (26.8) | 649 (36.3) | 114 (6.4) | 1787 (100) |
| Coping with external disturbances (such as nearby conversations) | 103 (5.8) | 350 (19.6) | 503 (28.2) | 565 (31.6) | 265 (14.8) | 1786 (100) |

#### Industry breakdown

The respondents selected the ownership of their company/organization from five options (governmental, county council/regional, municipal, private, non-profit organization/foundation). They also reported which industry the company/organization’s main activity belonged to by selecting one of the 16 categories of the Swedish Standard Industrial Classification (SNI) from 2007, which is based on the recommended standards from the European Union, NACE Rev.2 [20].

Different ways to categorize labour market sectors may result in different assumptions about managers’ opportunities to understand employee work capacity. The present study adopted an explorative approach rather than testing such assumptions. We used three different classifications to get as broad a picture as possible.

Classification 1: Ownership. Respondents were classified into the following groups: governmental, county council/regional and municipal, and private and non-profit organization/foundation based on their responses to the question about ownership in the survey.

Classification 2: Work object. The main work object (data, things or people) was used to classify respondents into the following groups: white-collar work (data), blue-collar work (things), pink-collar work (people) and other work (Table 2) [21]. The classification was done using the 16 categories of the SNI. The division into these four categories is a crude division because most occupations and jobs within an industry contain tasks related to all three objects. However, the significance of the main work object for job content has been recognized in research for decades, as well as the generalizations it may foster [22].

**Table 2.**
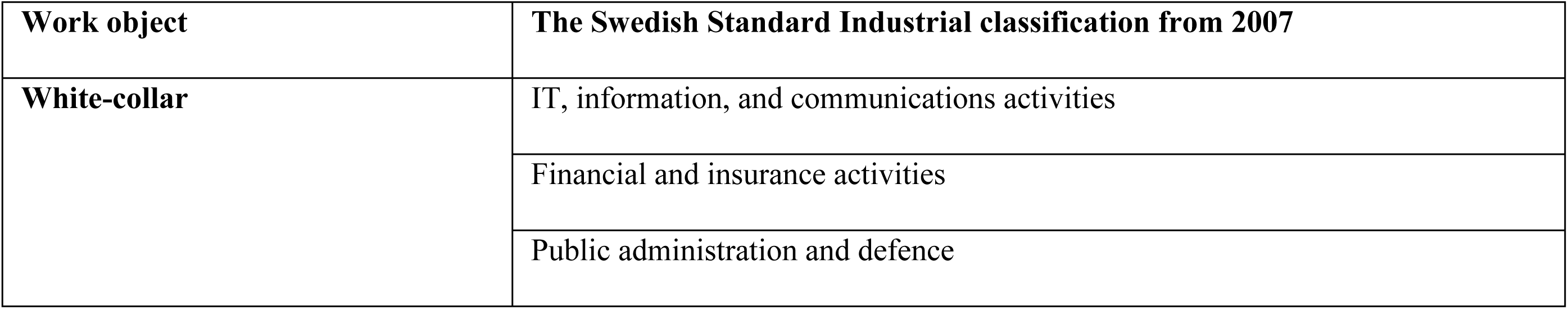
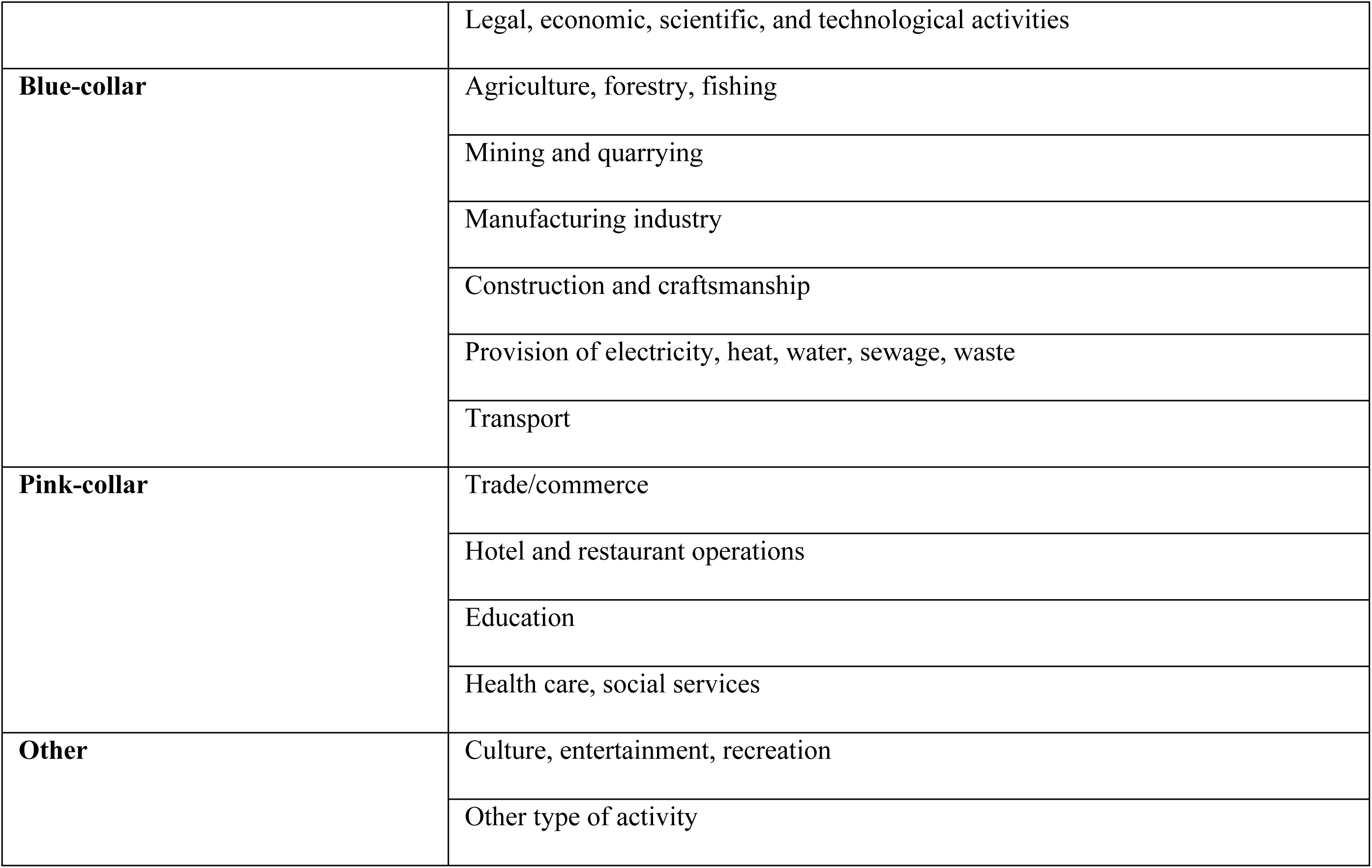
Classification of industries for classification 2: work object.

| Work object | The Swedish Standard Industrial classification from 2007 |
| --- | --- |
| White-collar | IT, information, and communications activities |
|  | Financial and insurance activities |
|  | Public administration and defence |
|  | Legal, economic, scientific, and technological activities |
| **Blue-collar** | Agriculture, forestry, fishing |
|  | Mining and quarrying |
|  | Manufacturing industry |
|  | Construction and craftsmanship |
|  | Provision of electricity, heat, water, sewage, waste |
|  | Transport |
| **Pink-collar** | Trade/commerce |
|  | Hotel and restaurant operations |
|  | Education |
|  | Health care, social services |
| **Other** | Culture, entertainment, recreation |
|  | Other type of activity |

Classification 3: Work object + gender composition. As a more fine-grained way of breaking down industries, we added the gender composition of the industry (female-dominated, male-dominated or gender-mixed). Following Cerdas et al. [23], both the main work object and the proportion of men and women employed in the industry were considered to classify the respondents into two male-dominated industries handling things (goods and energy production and machinery operations), three gender-mixed industries handling things, data and people (labour-intensive services, knowledge-intensive services, and public administration) and two female-dominated industries handling people (education and health and social care) (Table 3). The answer option “other type of activity” was excluded in this classification.

**Table 3.** Classification of industries for classification 3: Work object + gender composition.

| Work object + gender composition | The Swedish Standard Industrial classification from 2007<sup>a</sup> |
| --- | --- |
| **Machinery operations** | Agriculture, forestry, fishing |
|  | Construction and craftsmanship |
|  | Transport |
| **Goods and energy production** | Mining and quarrying |
|  | Manufacturing industry |
|  | Provision of electricity, heat, water, sewage, waste |
| **Labour-intensive services** | Trade/commerce |
|  | Hotel and restaurant operations |
|  | Culture, entertainment, recreation |
| **Knowledge-intensive services** | IT, information and communications activities |
|  | Financial and insurance activities |
|  | Legal, economic, scientific, and technological activities |
| **Education** | Education |
| **Health and social care** | Health care, social services |
| **Public administration** | Public administration and defence |

<sup>a</sup>The category, other type of activity, was excluded.

#### Size of the organization and span of control

It could be argued that the prospects for managers to rate how work capacity is affected are, to some extent, dependent on contextual factors, such as organizational size [24, 25] and span of control [26], i.e. the number of employees per manager. With large numbers of employees, it is plausible that managers have less insight into everyone’s work capacity. Organizational size was measured by the number of staff, collapsed into two categories: small and medium-sized organizations (0–250 staff members) and large organizations (>250 staff members) [27]. The span of control was collapsed into two groups: 0–30 and ≥31 employees per manager. In Sweden, 30 employees per manager is the average in the health and social sector, the sector with the largest span of control on the labour market [28]; more than 31 employees per manager is thus considered to be a large span of control. Descriptive statistics of the study sample, including demographics, are shown in Table 4.

**Table 4.**
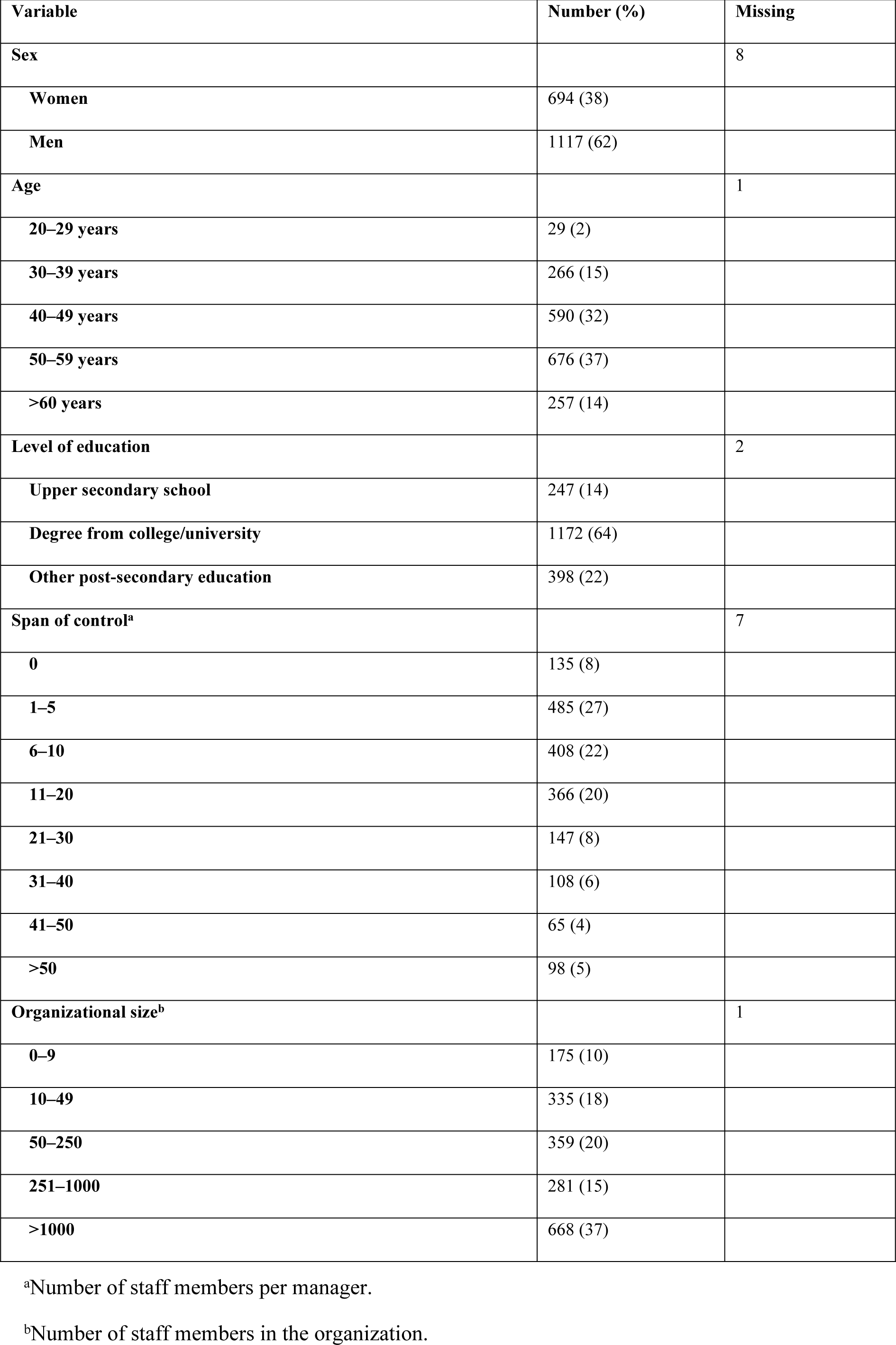
Descriptive statistics of the study sample (N = 1819).

| Variable | Number (%) | Missing |
| --- | --- | --- |
| **Sex** |  | 8 |
| **Women** | 694 (38) |  |
| **Men** | 1117 (62) |  |
| **Age** |  | 1 |
| **20–29 years** | 29 (2) |  |
| **30–39 years** | 266 (15) |  |
| **40–49 years** | 590 (32) |  |
| **50–59 years** | 676 (37) |  |
| **>60 years** | 257 (14) |  |
| **Level of education** |  | 2 |
| **Upper secondary school** | 247 (14) |  |
| **Degree from college/university** | 1172 (64) |  |
| **Other post-secondary education** | 398 (22) |  |
| **Span of control<sup>a</sup>** |  | 7 |
| **0** | 135 (8) |  |
| **1–5** | 485 (27) |  |
| **6–10** | 408 (22) |  |
| **11–20** | 366 (20) |  |
| **21–30** | 147 (8) |  |
| **31–40** | 108 (6) |  |
| **41–50** | 65 (4) |  |
| **>50** | 98 (5) |  |
| **Organizational size<sup>b</sup>** |  | 1 |
| **0–9** | 175 (10) |  |
| **10–49** | 335 (18) |  |
| **50–250** | 359 (20) |  |
| **251–1000** | 281 (15) |  |
| **>1000** | 668 (37) |  |

<sup>b</sup>Number of staff members in the organization.

### Data analysis

Descriptive statistics were applied to address the first research question. Differences in proportions between categories of industries regarding managers’ experience of one versus several employees with CMDs were explored using the chi-squared test. To address the second research question, three separate multivariate analyses of covariance (MANCOVAs), one per industry classification, were performed to investigate whether the industry was associated with managers’ ratings of how employees’ work capacity was affected by a CMD, controlling for organizational size and the managers’ span of control. County councils/regional and municipal, pink collar and health and social care were the reference categories in the MANCOVAs. The MANCOVAs were also performed using the full variance of the covariate, but because the results from these analyses did not differ from the analysis with dichotomized covariates, only the latter are presented in the results. All analysis was performed using IBM-SPSS software, version 26 (IBM, Armonk, NY, USA). The level of significance was set at <0.05.

## Results

### Managers’ experience of employees with CMDs

Managers’ experiences of employees with CMDs in different parts of the labour market where the industry is viewed according to ownership, work object and work object + gender composition are presented in Table 5. Table 5 also shows the results of the chi-squared tests of differences in managers’ experiences of staff members with a CMD, adjusted for the size of the organization and the span of control.

**Table 5.** Managers’ experience of employees with CMDs per industry.

| Industry breakdown | Experience of one employee with a CMD, n (%) | Experience of several employees with a CMD, n (%) | p value derived from the chi-squared test<sup>a</sup> |
| --- | --- | --- | --- |
| Ownership<sup>b</sup> |  |  |  |
| Governmental | 119 (43.4) | 155 (56.6) | <0.001 |
| Non-profit | 58 (43.6) | 75 (56.4) |  |
| Private | 543 (57.8) | 397 (42.2) |  |
| Municipal+ county councils | 207 (43.9) | 264 (56.1) |  |
| Work object |  |  |  |
| Other | 141 (48.8) | 148 (51.2) | <0.001 |
| Blue | 279 (52.2) | 255 (47.8) |  |
| White | 215 (60.2) | 142 (39.8) |  |
| Pink | 292 (51.0) | 892 (49.0) |  |
| Work object + gender composition<sup>c</sup> |  |  |  |
| Machinery operations | 89 (58.2) | 64 (41.8) | <0.001 |
| Goods and energy production | 126 (61.8) | 78 (38.2) |  |
| Labour-intensive services | 110 (52.1) | 101 (47.9) |  |
| Knowledge intense services | 181 (55.0) | 148 (45.0) |  |
| Education | 101 (43.7) | 130 (56.3) |  |
| Public administration | 98 (47.8) | 107 (52.2) |  |
| Health and social care | 123 (43.3) | 161 (56.7) |  |

<sup>a</sup>Chi-squared test regarding differences in proportions between classification categories of industries regarding managers' experience of one versus several employees with CMD controlled for the number of staff in the organization and number of employees per manager.
<sup>b</sup>One missing case,
<sup>c</sup>161 cases were excluded (the "other" category was excluded).

There was a difference between industries in the proportion (%) of managers with experiences of one versus several staff members with CMDs, irrespective of the classification. When the industries were defined according to ownership, the difference was due to one deviating industry; private companies had a lower proportion (42%) of managers who had experienced several staff members with CMDs, compared with the managers in other industries (56%–57%). We found a similar pattern when the industries were categorized in terms of the main work object; in white-collar industries, 40% of the managers had several employees with CMDs, whereas this proportion in other industries varied between 48% and 51%. For the third type of industry breakdown, work object + gender composition, the highest proportion of managers with several experiences of CMDs was found among health and social care and education (57%), followed by managers in public administration (52%), labour-intensive services (48%), knowledge-intensive services (45%), machinery operations (42%) and goods and energy production (38%).

### Managers’ ratings by industry classification

Three MANCOVAs were performed to assess the managers’ ratings of how work capacity was affected by CMDs (one for each industry breakdown: ownership, work object and work object + gender composition) with organizational size and span of control as covariates, and with task-oriented work capacity and relational work capacity as dependent variables. When industry was classified according to ownership, the results were F(6, 3518)=1.458, p=0.189, Wilks’ Λ=0.995, partial η^2^=0.002. For work object, the analysis resulted in F(6, 3518)=1.067, p=0.380, Wilks’ Λ=0.996, partial η^2^=0.002, and for work object + gender composition, F(12, 3118)=1.582, p=0.089, Wilks’ Λ=0.988, partial η^2^=0.006. Thus, no statistical differences could be found between industries on the combined measures of managers’ ratings of how work capacity is affected by a CMD, regardless of how the industry is classified.

## Discussion

We found that the proportion of managers who had experienced several staff members with CMDs was higher in industries where many women work: municipalities and counties, pink collar and education and health and social care. Differences attenuated when the classification of industries was based on work object. However, in contrast to our assumption, we did not find differences in how managers rated work capacity in employees with a CMD across industries.

### Managers’ experience of employees with CMDs

The finding that managers in industries where many women work encounter employees with CMDs to a greater degree than managers in other industries is in line with earlier research in which outcomes such as prescription of psychotropic drugs and psychiatric care [29], SA [9], and disability pension [30] with a CMD were found to be more prominent in female-dominated industries. Two systematic reviews have suggested a causal link between a detrimental psychosocial work environment and depression and burnout [31, 32]. Both reviews concluded that the negative consequences of work seem to affect women and men working in these environments in similar ways. This suggests that more women are affected because they often work in environments where factors that affect mental health negatively (e.g. job insecurity, high demands, high workload, low job control and low reward) are common [32].

Another reason for these differences is a possible underreporting of experiences. Managers are largely dependent on employee disclosure. Disclosure is a problematic issue, and workers avoid disclosure due to fear of stigmatization [33, 34]. In a recent qualitative study, managers reported that a masculine culture with traditional male ideals of being strong and enduring may contribute to stigma and hamper disclosure [35]. In another study, we found a difference in negative attitudes to depression between female and male managers; the proportion of women with negative attitudes was significantly lower than among men [36]. These two studies support the assumption that CMDs may be underestimated in male-dominated sectors. However, having employees with CMDs seems to be a rare experience for most managers; of the total study population (3358), 73% reported none or only one event in the last 2 years. The assumption regarding underreporting in male-dominated sectors must be confirmed in future studies.

### Managers’ ratings of how work capacity is affected by CMDs

Our next research question concerned differences in how managers in different industries in the labour market rate their employees’ work capacity in relation to CMDs. We know that organizational size and span of control vary between industries and that this can affect the managers’ possibilities to be attentive to their employees’ needs; therefore, we adjusted the analysis for these covariates. However, no main effect of industry, regardless of how the industry was measured, was found. The finding was a bit surprising because SA, which is a result of reduced work capacity, varies between industries. We suggest some possible explanations. First, the finding may imply that CMDs affect work capacity in similar ways across industries, at least from the managers’ perspective. Second, the nine items we used to illustrate possible effects of CMDs on work capacity did not correctly reflect different managers’ perspectives; other effects might be more visible or important to managers depending on the sector. Third, work capacity is a complex phenomenon that might be difficult to separate from symptoms such as fatigue, cognitive impairment, emotional instability, etc.

Our first possible explanation was that CMDs may affect work capacity in similar ways, irrespective of industry. This might be due to an alignment of work tasks in different sectors related to automation of repeated and/or heavy work tasks and digitalization. From theory, we know that work capacity is dependent on the environment, the work tasks, the demands and the resources of different workplaces [15]. If we narrow the perspective down to the individual employee, experiences of symptoms vary and affect capacity in different ways [16, 37, 38]. We therefore consider it unlikely that the absence of differences between sectors is due to alignment of work tasks or, for that matter, similar expressions of CMD-related work capacity, independent of the work situation. Rather, we acknowledge that the chosen macro level of comparison was too comprehensive to find variations in manager ratings. We suggest that future studies should be performed at lower structural levels to allow more specific comparisons. The challenge for that kind of study might be to find a large enough number for the analysis because employees with CMDs according to this study are a rare experience for managers.

Our second explanation concerns the nine items used in the survey to measure how managers rate work capacity in employees with CMDs. The items capture two aspects of work capacity: one on the relational aspects of work and one that was more task-oriented. It could be argued that this division of work capacity may be too fine-tuned for the managers. It is plausible that managers have a more general conception of work capacity among their employees. They may be able to conceive that work capacity is reduced, without being able to identify specific aspects that are problematic. However, in an earlier focus group study, managers from a variety of male- and female-dominated sectors were able to describe different components of their employees’ work capacity [39]. Thus, we have no particular reason to believe that the failure to find differences between industries was related to managers’ inability to identify specific aspects of reduced work capacity in their employees.

Although some managers may find it difficult to conceive different components of a reduced work capacity, we assume that they are randomly distributed over the industries. However, the items might be difficult to understand or to transfer into practical work experiences.

The third explanation for the lack of differences between sectors is the complexity of work capacity as a phenomenon. The complexity has been highlighted in several studies on clinical experience, where physicians state that the assessment of work capacity is difficult due to individual variations in symptoms and symptom management within similar groups of disorders [40, 41]. Also, persons affected by CMDs and reduced work capacity find it difficult to articulate their reduced work capacity. A qualitative study testing the Capacity Note, a communication facilitator in Swedish primary health care, found that patients thought it was helpful to get more precise wording of their experiences [42]. It made it easier to explain to their managers what they experienced as difficult in their work situation. Thus, the capacity to work concept might be less familiar to managers and employees with CMDs.

### Methodological considerations

To the best of our knowledge, this is the first study to consider managers’ perceptions of the work capacity of employees with CMDs across the labour market. One strength is that we tested three ways to classify managers according to industry, based on three logics, or theories (ownership, work object and work object + gender composition), with four, four and seven categories, respectively. Even though the sample was large, a single classification with seven categories could have hidden differences between industries. This risk of too many categories hiding possible differences was thus avoided.

To differentiate between managers with little and more extensive experience, we compared managers who had experienced one employee with a CMD with those who had experienced several staff members with CMDs in the past 2 years. The difference between the two categories can be disputed; the category “several staff members” includes both managers with extensive experience and those with just one more experience than the managers in the first category. Even with this crude division of managers’ experience, we managed to find differences between industries in the proportion of managers who had experienced employees with CMDs.

We were not in control of what kind of case the managers thought of when they were asked about their experience of employees with CMDs. Some might have thought about severe cases where employees were sick-listed and incapable of work, and others about cases when employees simply shared their health-related worries. The terms “depression and anxiety disorders” were used repeatedly throughout the survey to guide the managers’ understanding. However, we cannot be sure how the managers defined “employees with CMDs”. Managers from health care and social services may be better prepared due to formal training and experience to understand this concept than managers from other sectors. However, they did not rate work capacity differently. Furthermore, rating of work capacity among those with experience of several employees meant they had to rate an overall or general perception, which limits a nuanced rating.

### Implications

Managers rate capacity to work in employees with CMDs similarly, irrespective of the industrial sector. If the findings had supported our assumption about industry differences, tailored training programmes would have been warranted. However, the findings imply that the industrial context does not need to be considered in measures taken to support managers in the challenging management task of meeting employees with CMDs. The important issue is that managers across the labour market do receive support so that they can function as important actors in reducing work-related risk factors for CMDs and supporting sick-listed staff members to get back to work.

## Conclusion

This study aimed to investigate whether there were differences among managers in the various industries in the Swedish labour market regarding experiences of CMDs in employees, and how managers rated the effect of CMDs on employees’ work capacity. It was found that the proportion of managers who had experienced several staff members with CMDs in the past 2 years was higher in industries where many women work, regardless of whether the industry was classified according to ownership, work object or work object in combination with gender composition. However, no difference could be found in how managers from different industries rated how the employees’ work capacity was affected by CMDs.

## Data Availability

The data used for this study are archived at the Laboratory of Opinion Research (LORE) at the University of Gothenburg and can be obtained by contacting LORE at

## Supporting information

**S1 Table.** The results of the principal component factor analysis with the rotated component matrix.

| Variable | Was the employee's work capacity affected regarding | Component |  |
| --- | --- | --- | --- |
|  |  | 1 | 2 |
| 1 | Getting started on work tasks | 0.792 | 0.085 |
| 2 | Being able to prioritize among work tasks | 0.770 | 0.215 |
| 3 | Interacting with other people | 0.223 | 0.684 |
| 4 | Being able to concentrate | 0.701 | 0.361 |
| 5 | Remembering | 0.637 | 0.363 |
| 6 | Maintaining the work pace | 0.714 | 0.177 |
| 7 | Participating in social work environments (such as coffee breaks) | 0.106 | 0.768 |
| 8 | Keeping calm and not be upset | 0.184 | 0.679 |
| 9 | Coping with external disturbances (such as nearby conversations) | 0.323 | 0.640 |

## Notes

### Competing Interest Statement

The authors have declared no competing interest.

### Author Declarations

The project was approved by the Regional Ethics Review Board in Gothenburg (Dnr: 165-17). Written consent to participate were provided by all informants.

